# Risk factors for antibiotic misuse for cellulitis in the emergency department through the SARS-CoV-2 pandemic: A single institute retrospective study

**DOI:** 10.1101/2025.10.22.25338374

**Authors:** Maya R. Navarro, Mingyu Qin, Ziyou Ren

## Abstract

Cellulitis has a large burden on emergency departments (EDs) annually in the United States both in case volume and recurrence, causing antibiotic waste and poorer patient outcomes. To identify medical and demographic risk factors independently associated with antibiotic overprescription for cellulitis in the ED, we employed a retrospective cohort study utilizing electronic health records on patients with cellulitis-like symptoms in the ED from 2016 to 2023. Demographics, such as race, ethnicity, insurance status, and comorbidities were matched and reviewed alongside patient medical records to determine the over-prescribed cohort. A total of 1,283 (9%) of 13,530 patients were over-administered antibiotics for cellulitis-like-symptoms in the ED. Multivariate analysis showed that white, non-Latinx heritage (Relative risk (RR)=1.60; p<0.0001) and private insurance status (RR= 1.58; p<0.0001) patients were the most at risk for inappropriate antibiotic administration. Patients with chronic pulmonary disease (RR= 1.39; p<0.0001) had the strongest independent medical association with antibiotic over-administration. While this is a retrospective study limited to one institution’s patient population, our results demonstrate that antibiotic administration practices for cellulitis in the ED is significantly associated with patient race and insurance status, reflecting current health disparities.

## Introduction

Cellulitis is an acute, infectious inflammation of the dermis and subcutaneous tissue [26] which accounts for an estimated 14.5 million cases annually in emergency departments (EDs) across the United States [21]. Up to 85% of cases of cellulitis are non-culturable [3] and up to 45% of cases per year recur in the same limb [28], suggesting immediate broad-spectrum antibiotic administration in the ED for patients presenting with classic symptoms of cellulitis, such as erythema and edema accompanied by fever and malaise, may be inappropriate [26]. Common inflammatory dermatoses that mimic cellulitis lead to a misdiagnosis rate of cellulitis in non-dermatology clinic settings ranging from 30-60% [4, 16, 29] and the administration of empirical antibiotics and/or unnecessary hospitalization [12]. Recent efforts to quantify this waste show that the over-administration of broad spectrum antibiotics for cellulitis-like symptoms in the ED, at best, presents EDs with an estimated $900,000 annually wasted in antibiotics [23], and, at worst, may contribute to the recent surge in skin and soft tissue infections with antibiotic-resistant bacteria [9]. Further compounding the need to address antibiotic misappropriation in the ED is the increasing prevalence of multimorbidity– defined as living with two or more chronic medical conditions– which has complicated ED workflows, contributed to higher rates of ED readmission, longer ED stays, and overall worse patient outcomes [30].

Although EDs across the United States saw declines in usage during Q2 2020 when the COVID-19 pandemic began, adult ED visits for all demographic factors have since rebounded to or have since increased compared to pre-pandemic levels [22, 25]. However, at the time of writing, there is no literature which attempts to categorize rates of ED utilization or of antibiotic misappropriation for cellulitis since the SARS-Cov-2 pandemic.

Prior studies have sought to identify cases of cellulitis that could be managed without hospitalization or antibiotics, providing a framework for our “over-prescribed” cohort criteria. For instance, Kroshinsky et al. (2007) and Lazzarini et al. (2005) demonstrated that normal inflammatory markers (e.g., WBC count, CRP) correlate with milder cellulitis cases unlikely to require systemic antibiotics [12, 13]. Similarly, Cutler et al. (2023) and Nightingale et al. (2023) found that misdiagnosis-driven antibiotic overuse often manifests as short-course prescriptions (<7 days) or rapid ED returns (<30 days), reinforcing our inclusion of these criteria [4, 16]. These findings align with clinical guidelines emphasizing judicious antibiotic use to mitigate resistance risks [6, 21] and reduce unnecessary hospitalizations [9, 29]. Thus, our cohort definitions reflect evidence-based thresholds for distinguishing low-severity cellulitis amenable to conservative management.

Therefore, the primary objective of this study was to identify both medical and demographic risk factors independently associated with the overprescription of antibiotics to patients with cellulitis-like symptoms in the ED. We also aimed to evaluate ED utilization trends and multimorbidity burden for cellulitis patients at our institution, specifically focusing on changes since the SARS-CoV-2 pandemic.

## Methods

### Data Source

The Northwestern Medicine Enterprise Data Warehouse (NMEDW) is a large electronic medical record repository which stores data on over 6.6 million distinct patients in the Northwestern Medicine system [14]. The NMEDW contains billing claims data from as early as 1998 and electronic health records from as early as 2002, however, data entry on the NMEDW platform was standardized starting in 2007. To maximize the quality of the data collected through the NMEDW query, only patient charts from 2016 to 2023 were considered. Patients that did not have a complete blood count (CBC) with a differential attached to their progress note, or which otherwise had incomplete data pulled from the NMEDW, were excluded from further analysis.

### Study Design

We conducted a retrospective cohort study of patients presenting to emergency departments within the Northwestern Memorial Healthcare System between January 2016 and December 2023. Patient records were obtained through the Northwestern Medicine Enterprise Data Warehouse (NMEDW) and screened for a primary diagnosis of cellulitis using ICD-10 codes (L03.90, L03.01, L03.03, L03.11, L03.211, L03.221, L03.31, or L03.81). We excluded patients under 18 years of age, pregnant patients (ICD-10 codes O00-O9A), and those not evaluated in an emergency department setting. Additional exclusion criteria included patients with clinical conditions likely to influence antibiotic prescribing decisions, as described in prior studies [16, 29]. Specifically, we excluded patients with recent surgical procedures within 30 days of ED presentation (ICD-10 codes Y83.1-Y83.9, Y84.1-Y84.9, Y62.1-Y62.9, or Y65.1-Y65.9), documented osteomyelitis (ICD-10 codes M86.0-M86.9), diabetic foot ulcers (ICD-10 codes E11.621, E11.622, or E11.628), or indwelling medical devices (ICD-10 codes T85.698A or T83.9XXA).

Patients were classified into the over-prescribed cohort if they met any of three predefined criteria established in prior research. First, patients with normal white blood cell counts and C-reactive protein levels measured prior to antibiotic administration were included, based on evidence that these laboratory values correlate with low-severity cases that may not require antimicrobial therapy [12, 13]. Second, patients who returned to the emergency department with recurrent cellulitis within 30 days of their initial visit were included, as this pattern has been associated with misdiagnosis in previous studies [4, 16]. Third, patients who received antibiotic courses of seven days or less were included, as shorter durations have been linked to inappropriate prescribing for non-bacterial conditions [4, 24]. The specific antibiotics considered included oral formulations (trimethoprim-sulfamethoxazole, amoxicillin, doxycycline, linezolid, dicloxacillin, flucloxacillin, cephalexin, cefadroxil, or clindamycin) and intravenous formulations (vancomycin, cefazolin, cefepime, daptomycin, meropenem, nafcillin, oxacillin, or flucloxacillin).

To ensure accurate classification, all patients identified through these automated criteria underwent manual chart review by trained research staff. This validation step helped prevent misclassification of patients who may have appropriately received antibiotics for concurrent conditions. Our approach aligns with current antimicrobial stewardship principles aimed at reducing unnecessary antibiotic use while maintaining patient safety [6, 11]. The study protocol was reviewed and approved by the Northwestern University Institutional Review Board (approval #STU00218213).

### Statistical Analysis

R software (version 3.0.2) was used to conduct the statistical analysis. Logistics regression models using the STATS package (version 4.6.0) were used to determine relative risk (RR), 95% confidence intervals (CIs), and p-values of demographic-specific contribution to antibiotic misappropriation. MedCalc for Windows, version 23.2.1 (MedCalc Software, Ostend, Belgium) was used to validate statistical calculations. A 2-way ANOVA was run on the variables of race/ethnicity and insurance status to determine if the relative risk of antibiotic overprescription for cellulitis was both significant and independent.

## Results

Out of the 13,530 unique patients meeting inclusion criteria, ~9% (n= 1,283) of patients were determined to be over-prescribed antibiotics in the ED for cellulitis-like symptoms. Of the over-prescribed cohort, 91.1% had a within reference range white blood cell count on CBC with differential (n=1169), 23.3% had a within reference range CRP (n=298), and 69.1% were readmitted to the ED within 30 days of their first cellulitis diagnosis (n=886). The prevalence of antibiotic misappropriation increased overall by 75% since 2016 and did not significantly change during the SARS-Cov-2 pandemic as rates of emergency department utilization generally increased across all demographics starting in 2021 (Figure 1).

**Fig. 1.**
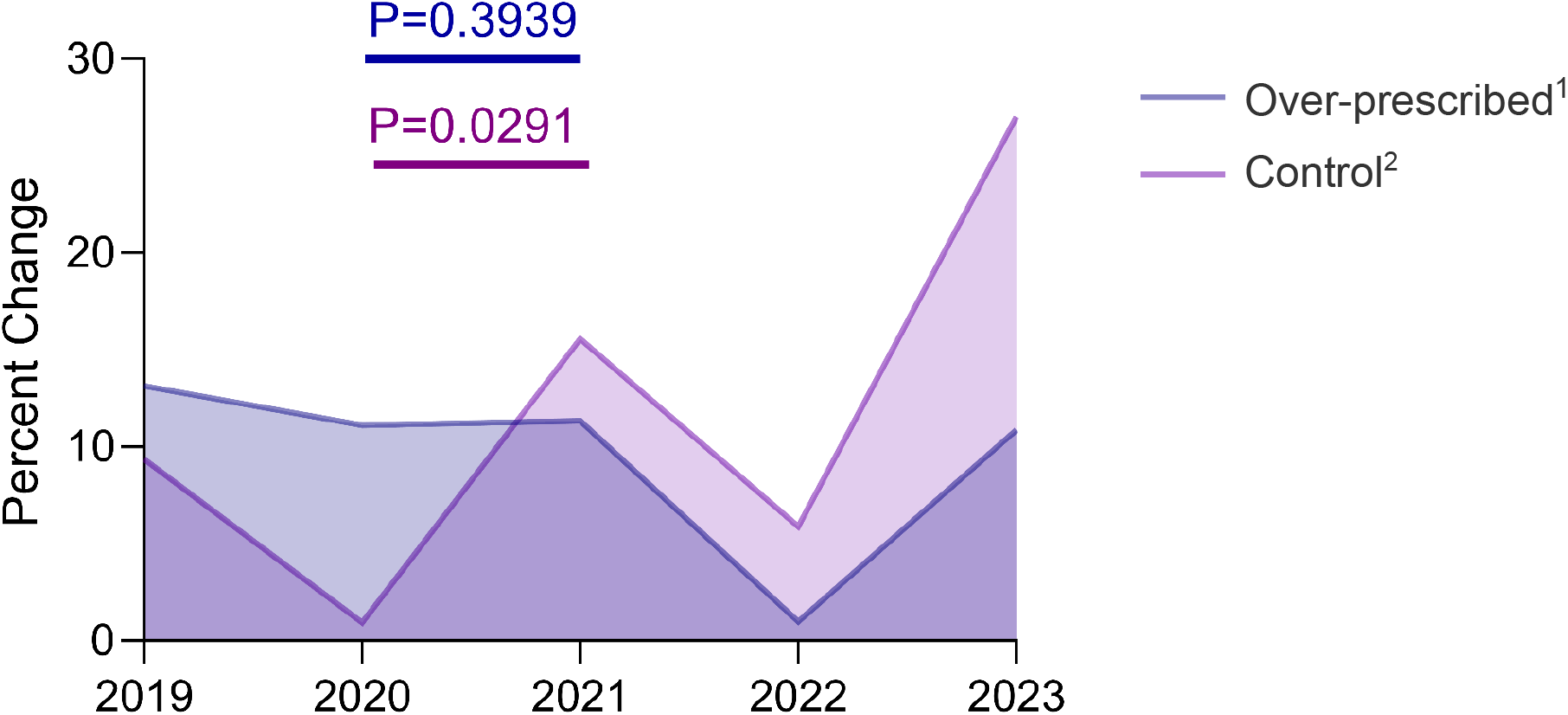
Overall emergency department utilization trends from 2019-2023 for patients with cellulitis-like symptoms. ^1^ Percent change across patients over-administered antibiotics for their cellulitis-like symptoms in the ED did not significantly change (P=0.39) across pre- and post-pandemic cohorts. ^2^ Average percent change in ED utilization for cellulitis increased by 108% (p=0.029) between pre- and post-pandemic cohorts, irrespective of antibiotic misappropriation and other demographic factors.

Disparities for antibiotic overprescription appear in the yearly trends for patient race and insurance status, respectively. White patients were at a 1.6 times greater (CI 1.39-1.85; p<0.0001) risk of being over-prescribed antibiotics for cellulitis whereas Black patients have a RR of 0.41 (CI 0.32-0.54; p<0.0001) and Hispanic/Latinx patients have a RR of 0.66 (CI 0.54-0.80; p<0.0001) (Table 1). The most notable change as a result of the SARS-CoV-2 pandemic was observed among individuals identifying as White, who experienced a 131% increase in ED utilization (p=0.085) between pre- and post-pandemic cohorts, irrespective of antibiotic misappropriation status. In contrast, other racial and ethnic groups did not show statistically significant changes: Black individuals had a 19.4% increase (p=0.48), Hispanic/Latine individuals saw a 660% increase (p=0.38), and Asian individuals displayed a 961% decrease (p=0.58). Additionally, patients who declined to report their race or ethnicity exhibited a 581% increase (p=0.43) (Figure 2a). These findings suggest that the pandemic may have disproportionately impacted ED utilization for cellulitis among White patients, while changes in other groups were not significant.

**Table 1.**
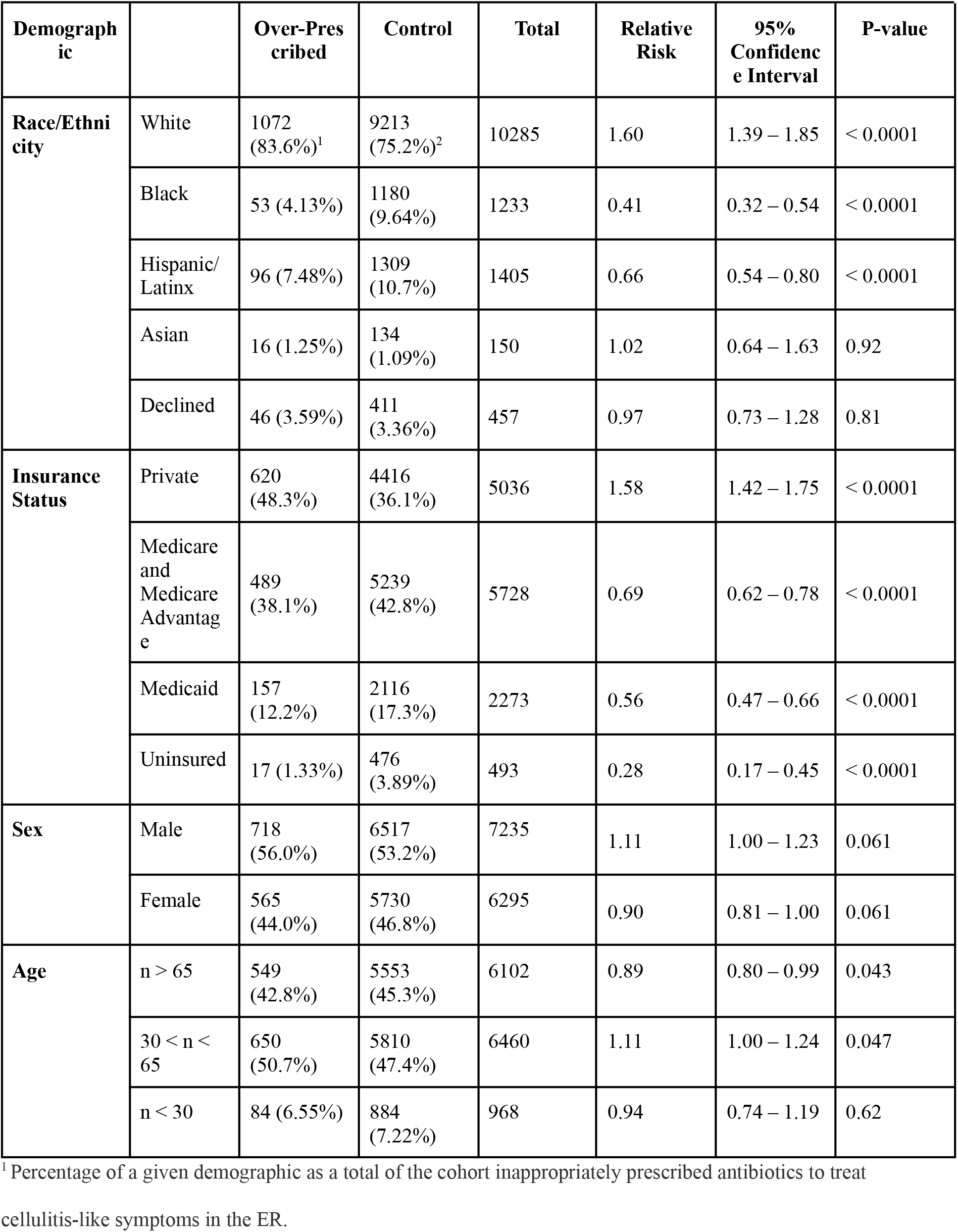
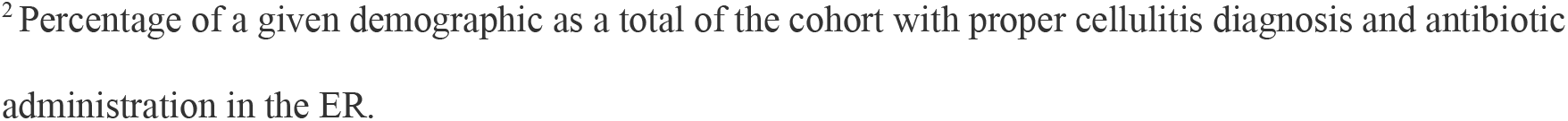
“Risk of antibiotic over-administration based on multiple sociodemographic factors.”

| **Demographic** |  | **Over-Prescribed** | **Control** | **Total** | **Relative Risk** | **95% Confidence Interval** | **P-value** |
| --- | --- | --- | --- | --- | --- | --- | --- |
| **Race/Ethnicity** | White | 1072<br>(83.6%)<sup>1</sup> | 9213<br>(75.2%)<sup>2</sup> | 10285 | 1.60 | 1.39 – 1.85 | < 0.0001 |
|  | Black | 53 (4.13%) | 1180<br>(9.64%) | 1233 | 0.41 | 0.32 – 0.54 | < 0.0001 |
|  | Hispanic/Latinx | 96 (7.48%) | 1309<br>(10.7%) | 1405 | 0.66 | 0.54 – 0.80 | < 0.0001 |
|  | Asian | 16 (1.25%) | 134<br>(1.09%) | 150 | 1.02 | 0.64 – 1.63 | 0.92 |
|  | Declined | 46 (3.59%) | 411<br>(3.36%) | 457 | 0.97 | 0.73 – 1.28 | 0.81 |
| **Insurance Status** | Private | 620<br>(48.3%) | 4416<br>(36.1%) | 5036 | 1.58 | 1.42 – 1.75 | < 0.0001 |
|  | Medicare and Medicare Advantage | 489<br>(38.1%) | 5239<br>(42.8%) | 5728 | 0.69 | 0.62 – 0.78 | < 0.0001 |
|  | Medicaid | 157<br>(12.2%) | 2116<br>(17.3%) | 2273 | 0.56 | 0.47 – 0.66 | < 0.0001 |
|  | Uninsured | 17 (1.33%) | 476<br>(3.89%) | 493 | 0.28 | 0.17 – 0.45 | < 0.0001 |
| **Sex** | Male | 718<br>(56.0%) | 6517<br>(53.2%) | 7235 | 1.11 | 1.00 – 1.23 | 0.061 |
|  | Female | 565<br>(44.0%) | 5730<br>(46.8%) | 6295 | 0.90 | 0.81 – 1.00 | 0.061 |
| **Age** | n > 65 | 549<br>(42.8%) | 5553<br>(45.3%) | 6102 | 0.89 | 0.80 – 0.99 | 0.043 |
|  | 30 < n < 65 | 650<br>(50.7%) | 5810<br>(47.4%) | 6460 | 1.11 | 1.00 – 1.24 | 0.047 |
|  | n < 30 | 84 (6.55%) | 884<br>(7.22%) | 968 | 0.94 | 0.74 – 1.19 | 0.62 |

<sup>1</sup> Percentage of a given demographic as a total of the cohort inappropriately prescribed antibiotics to treat
cellulitis-like symptoms in the ER.
<sup>2</sup>Percentage of a given demographic as a total of the cohort with proper cellulitis diagnosis and antibiotic administration in the ER.

**Fig. 2.**
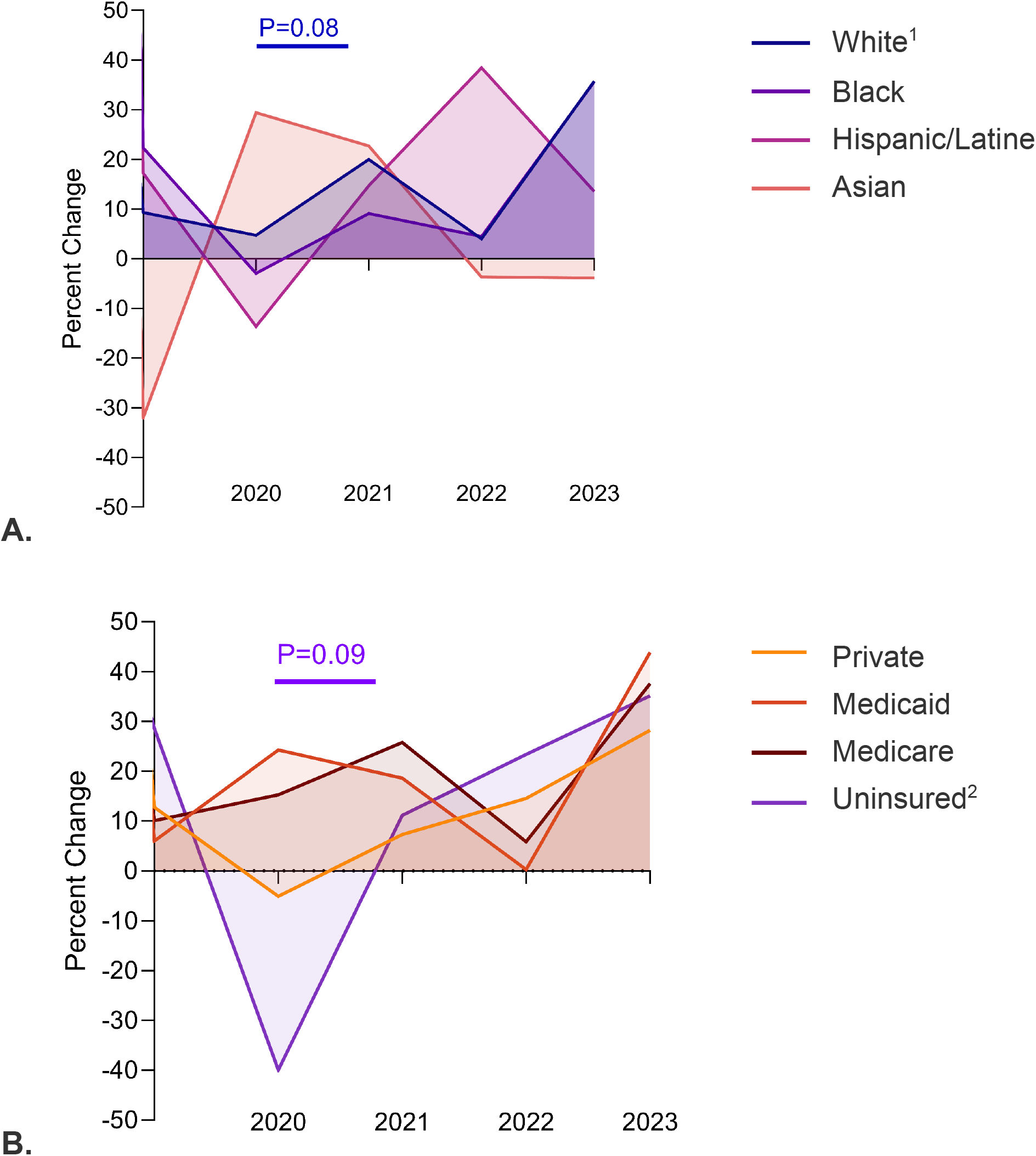
Emergency department utilization demographic trends from 2019-2023 for cellulitis a. Sorted by patient race and ethnicity. b. Sorted by patient insurance status at time of initial cellulitis diagnosis ^**1**^ Average percent change in ED utilization for cellulitis increased by 131% (p=0.085) between individuals identifying as white between pre- and post- pandemic cohorts, irrespective of antibiotic misappropriation status. No other races saw a significant difference in percent changes (Black: +19.4%, p=0.48; Hispanic: +660%, p=0.38; Asian: −961%, p=0.58 Declined: +581%, p=0.43). ^**2**^ Average percent change in ED utilization for cellulitis decreased by 229% between uninsured individuals in pre- and post-pandemic cohorts, regardless of antibiotic misappropriation status. No other patient insurance statuses exhibited significant percent change differences in ED utilization between pre- and post-pandemic periods (Privately insured: +195%, p=0.13; Medicaid recipients:+66.8%, p=0.43; Medicare recipients: +43.6%, p=0.59).

For insurance status, patients with a private insurer such as Blue Cross Blue Shield or a Commercial/Managed Care plan had a 1.58 times greater (CI 1.42-1.75; p<0.0001) risk of being over-prescribed antibiotics for cellulitis-like symptoms whereas patients with Medicare and Medicare advantage have a RR of 0.69 (CI 0.62-0.78; p<0.0001), patients with Medicaid and Medicaid replacement have a RR of 0.56 (CI 0.47-0.66; p<0.0001), and uninsured patients have a RR of 0.28 (CI 0.17-0.45; p<0.0001) (Table 1). Both privately insured and uninsured patients saw a significant decline in ED utilization for cellulitis-like symptoms during 2020 but uninsured patients decline in ED utilization was nearly one order of magnitude greater than that of privately insured patients (Figure 2b). Since 2021, rates of ED utilization have had a net positive percent change across patients of all insurance status (Figure 2b). 2-Way ANOVA results detailed that race/ethnicity of a patient (p=0.0002) and insurance status (p=0.0003) independently have significant effects on the risk of antibiotic misappropriation in the ED for cellulitis-like symptoms but there was no significant interaction (p=0.282) between these two key demographic features.

61% (n=779) of patients over-prescribed antibiotics had multimorbidity status at diagnosis, compared to 47% (n=6325) in the controls. The most common comorbidities from both the over-prescribed cohort and controls were diabetes (n=4247), chronic pulmonary disease (n=3261), and long term drug therapy (n=2622). Comorbidities most strongly associated with over-prescription were rheumatic disease (n=712, RR=1.65; p<0.0001; CI=1.35-2.03), peptic ulcer disease (n=326, RR=1.40; p=0.031; CI=1.03-1.91), chronic pulmonary disease (n=3261, RR=1.39; p<0.0001; CI=1.22-1.59), and malignancy (n=1727, RR=1.34; p=0.0005; CI=1.14-1.57) (Table 2). The RR of antibiotic over-prescription plateaued at an average of 1.10 if patients had more than two comorbidities and the number of comorbidities that a patient had at the time of cellulitis diagnosis did not significantly impact risk of antibiotic misappropriation (Figure S1).

**Table 2.** “Risk of antibiotic over-administration based on the total cohort’s most common comorbidities.”

| **Comorbidity** | **Over-Prescribed** | **Control** | **Total** | **Relative Risk** | **95% Confidence Interval** | **P-value** |
| --- | --- | --- | --- | --- | --- | --- |
| Diabetes | 355 | 3892 | 4247 | 0.96 | 0.83 – 1.10 | 0.52 |
| Chronic pulmonary disease | 397 | 2864 | 3261 | 1.39 | 1.22 – 1.59 | <0.0001 |
| Long term drug therapy | 220 | 2402 | 2622 | 0.96 | 0.82 – 1.13 | 0.61 |
| Congestive heart failure | 174 | 1908 | 2082 | 0.95 | 0.80 – 1.14 | 0.61 |
| Malignancy | 202 | 1525 | 1727 | 1.34 | 1.14 – 1.57 | 0.0005 |
| Cerebrovascular disease | 165 | 1404 | 1569 | 1.20 | 1.01 – 1.43 | 0.040 |
| Rheumatic disease | 103 | 609 | 712 | 1.65 | 1.35 – 2.03 | <0.0001 |
| Peptic ulcer disease | 40 | 286 | 326 | 1.40 | 1.03 – 1.91 | 0.031 |
| Moderate/severe liver disease | 5 | 134 | 139 | 0.41 | 0.17 – 0.98 | 0.043 |

## Discussion

We identified that race and ethnicity and insurance status independently have significant effects on the risk of antibiotic misappropriation for cellulitis in the ED. Uninsured patients, regardless of race or ethnicity, had the lowest risk of being over-administered antibiotics for their cellulitis symptoms, suggesting a general reluctance by ED providers to administer the same intensity of care as they would for insured patients presenting with similar symptoms. Additionally, Black and Hispanic/Latine patients have a lower relative risk of having antibiotics over-administered in spite of having higher rates in utilizing the ED to treat their cellulitis-like symptoms since the SARS-CoV-2 pandemic in comparison to white, non-Latine patients. This finding correlates with previous literature which demonstrated that Black and Hispanic/Latine patients were more likely to utilize the ED for ongoing medical conditions [18] yet were less likely to receive antibiotic prescription from the same clinician as compared to their white peers [6].

Our findings demonstrate that disparities in care intensity for cellulitis-like symptoms manifest asymmetrically across demographic groups. While white, privately insured patients face higher rates of antibiotic overprescription—reflecting inappropriate treatment—Black, Hispanic/Latine, and uninsured patients experience lower intervention rates despite comparable clinical need, suggesting under-treatment. This dichotomy underscores previous findings that health disparities operate bidirectionally: systemic biases lead to excessive care for some groups (e.g., unnecessary antibiotics for white patients) and insufficient care for others (e.g., withheld therapies for marginalized populations) [2, 10]. Crucially, both outcomes represent gaps in equitable care delivery, as overprescription harms patients through antibiotic-related adverse effects and antimicrobial resistance, while under-treatment risks untreated infections and increased ED revisits [7]. By reframing our conclusion to emphasize this dual dynamic, we clarify that disparities in “level and intensity of care” are not unidirectional but reflect broader systemic inequities in how race and insurance status influence clinical decision-making. These patterns align with national data showing that overprescription disparities often correlate with structural factors like implicit bias in pain perception and differential access to follow-up care, which disproportionately affect marginalized groups even when overtreatment metrics favor them [8, 15].

We hypothesize the reason for this racial disparity in antibiotic administration for cellulitis-like symptoms is twofold. First, current guidelines from the Accreditation Council for Graduate Medical Education (ACGME) do not detail curriculum plans to incorporate a fundamental overview of dermatologic conditions within emergency medicine residency programs, greatly stratifying the overall exposure that EM physicians receive in caring for skin disorders [12]. Second, the dearth of skin of color dermatology training across the field of medicine means that physicians in the ED may not have ever received training to recognize classic signs of cellulitis, particularly erythema, in skin of color [17]. Further studies should be performed to gauge emergency medicine provider comfortability with managing skin disorders in patients of color.

The clearest limitation in our findings is the limited sample size, as the data from the NMEDW pulls from the 12 emergency departments ran by one large healthcare institution located in Illinois, a state with a robust quarantine procedure during the SARS-Cov-2 pandemic yet variable vaccination rates across the state’s metropolitan and rural areas [5, 10]. As such, emergency department utilization trends and demographics discussed in this manuscript are only generalizable to patients serviced by the EDs located, at a maximum of 50 miles outside of the Chicagoland metropolitan area and does not capture emergency department utilization trends in more rural areas of the state in Central and Southern Illinois. Outside of geographic limitations, diagnoses listed on extracted patient data were made on clinical grounds from the team in the ED, which, at the time of writing, represents 121 individuals in the Department of Emergency Medicine faculty alone [1]. However, implicit and explicit bias from the clinical team supervising the ED may impact a patient receiving a cellulitis diagnosis in their medical record, thus providing an unintentionally misrepresentative patient population in the NMEDW. Moreover, while we were able to determine that patients with at least one comorbidity face the highest risk of inappropriately receiving antibiotics for cellulitis-like symptoms in the ED, we did not find that the overall degree of patient multimorbidity does not significantly influence antibiotic misappropriation risk.

In sum, our analysis of medical and demographic risk factors independently associated with antibiotic overprescription highlighted the impact of both a patient’s race and insurance status on the level and intensity of care they receive for their symptoms in the ED for cellulitis-like symptoms. White, privately insured patients are the most likely to receive excessive antibiotics in the ED to treat cellulitis-like symptoms and disparities in providing dermatologic care in the ED exist on the basis of the patient’s race and insurance status – reflecting the need for accessible skin of color curricula for ED providers.

## Data Availability

All data produced in the present study are available upon reasonable request to the authors.

## Statements and declarations

All authors have no disclosures/ no conflict of interest.

## Acknowledgements

This work was supported in part by the Northwestern Medicine Enterprise Data Warehouse. The authors would like to acknowledge EDW pilot data funding program and the Feinberg School of Medicine Summer Research Scholars Program for their financial and academic support in developing this project.

## Author Contributions

MRN and ZR contributed to the study conception and design. MQ extracted patient charts from the electronic data warehouse. MRN analyzed the data under the supervision of ZR. MRN and ZR wrote the manuscript. All authors have read and approved the final manuscript.

## SUPPLEMENTAL

**Supplemental Fig. 1.**
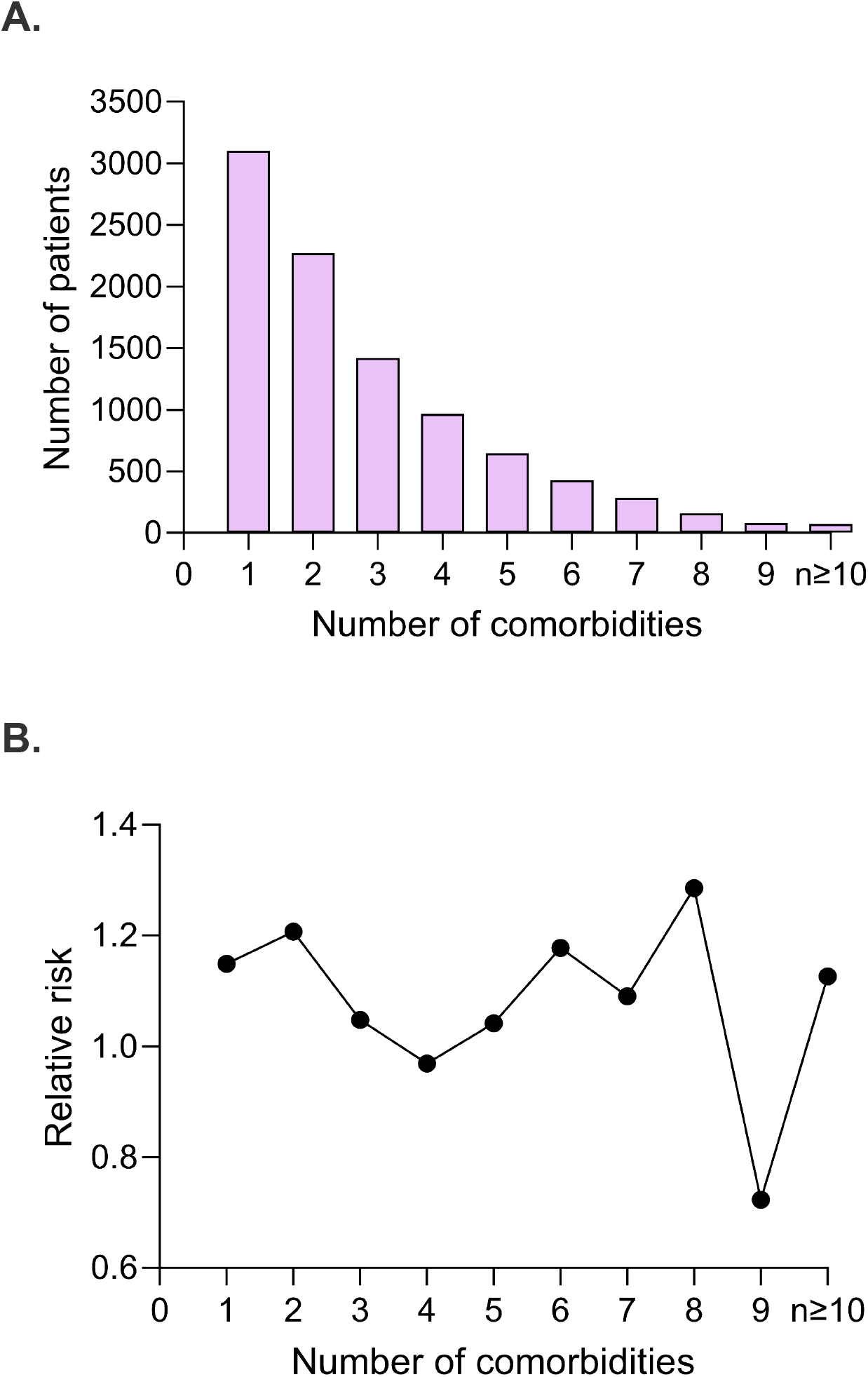
Degree of multimorbidity in patients in the ED with cellulitis-like symptoms a. Prevalence of multimorbidity. b. Relative risk versus degree of patient multimorbidity.

